# Prenatal exposure to extreme temperatures and neonatal health in Lausanne 1909 to 1912

**DOI:** 10.1101/2025.09.18.25336011

**Authors:** Ellen Hünerwadel, Katarina L Matthes, Noémie Letellier, Tarik Benmarhnia, Kaspar Staub, Mathilde Le Vu

## Abstract

**Background:** Extreme temperatures are increasingly recognized as risk factors for maternal and neonatal health, but historical evidence remains scarce. This study investigates the association between prenatal exposure to extreme heat and cold and neonatal health outcomes in Lausanne, Switzerland, during 1909–1913, including the exceptional 1911 heatwave.

**Data & Methods:** We digitized and linked daily minimum and maximum temperature data from the Champs-de-l’Air weather station with 2,000 maternity records from Lausanne’s hospital (1909–1913). Continuous outcomes included birth weight, gestational age, placenta weight, birth length, and head circumference; binary outcomes were low birth weight (LBW, <2,500g) and preterm birth (PTB, <37 weeks). Daily mean temperature exposure was averaged over whole pregnancies and trimesters. Associations were estimated using generalized linear models for continuous outcomes and Cox proportional hazards for binary outcomes, adjusting for maternal and seasonal covariates.

**Results:** Infants exposed to extreme heat (>90th percentile) or cold (<10th percentile) throughout pregnancy had consistently poorer outcomes compared with moderate exposure. Whole-pregnancy exposure to the 95th percentile temperature was associated with −211g lower mean birth weight and −1.1 weeks shorter gestation relative to median exposure. Cold exposure was linked to increased LBW risk but less strongly to shortened gestation. In unadjusted analyses, LBW prevalence was 25% under high-temperature exposure versus 10% at moderate levels; PTB prevalence was 26% versus 9%. Stillbirth and early neonatal mortality rates were also higher at temperature extremes. Effects were strongest for third-trimester exposure and more pronounced for heat than for cold.

**Conclusion:** This study provides rare historical evidence that both heat and cold during pregnancy adversely affected neonatal health in early 20th-century Switzerland, with heat exposure during the 1911 heatwave particularly detrimental. These findings underline that vulnerability of pregnant women and newborns to temperature extremes might be a long-standing phenomenon.

## 1. Introduction

Neonatal health can be influenced by many factors such as genetics and maternal health during pregnancy.^1^ Maternal stress also plays an important role, affecting fetal growth and other neonatal outcomes. Key indicators of neonatal health include fetal and neonatal mortality rates, gestational age, infant size, prevalence of congenital anomalies, and Apgar scores.^2^ These indicators are in turn associated with infant morbidity and mortality, as well as with later-life outcomes.^3^ For instance, it is well established that low birth weight (LBW) is associated with higher risks of morbidity and stunting in childhood,^4^ and an increased risk of chronic diseases in adult life.^5^

Environmental factors can also impede fetal development, such as extreme weather events and air pollution. Extreme heat exposure has been found to negatively affect fetal outcomes from stillbirth rates to birth weight, congenital anomalies and gestational age.^6–8^ The timing of heat exposure during pregnancy seems to be particularly important. The risk of being born preterm (<37 weeks of gestation) was higher for mothers exposed to extreme heat during the last week of gestation, with a dose-response relationship as the temperature and duration of the heat wave increased.^9^ A systematic analysis also reports that the risk of preterm birth and stillbirth were higher during the last week before birth.^10^ Exposure to cold temperatures is also associated with higher risks of preterm birth, stillbirth and a reduced fetal growth ^11^, including lower birth weight, placenta weight and birth length.^12–14^

With climate change, the frequency and intensity of heat waves is increasing,^15^ significantly threatening human health, including maternal and infant health.^16^ The World Health Organization (WHO) and the World Meteorological Organization (WMO) acknowledge the importance of national and local heat-health action plans in mitigating the effects of extreme temperatures on vulnerable populations, particularly in preventing heat-related illnesses and fatalities.^17,18^ While previous studies have identified vulnerable populations affected by extreme temperatures - especially infants and young children - research focusing specifically on the neonatal population remains limited.^19–21^ However, in environmental epidemiology, temperature related mortality has been an important issue for some time, especially in the course of climate change.^22^ Many regions of the world are affected by these negative impacts.^23^ Recently it was shown that in Switzerland, heat- and cold-related mortality corresponded to 2.4 and 77.0 deaths per 100,000 people annually, with an increasing tendency for heat, a big proportion being attributed to population aging as a main risk factor,^24^ and urban centers being particularly affected.^25^ It is likely that periods with abnormally high temperatures will continue to increase in the future, including in Switzerland.^26^

Notable examples include the 2003 European heatwave and the 1995 Chicago heatwave, both of which caused considerable public health impacts.^27,28^ However, heat waves have sporadically occurred much earlier in the 20^th^ century: in 1911, an important heat wave hit Central and Southern Europe, during which not only older people suffered, but also many infants.^29–31^ While increased infant and child mortality was observed during the 1911 European heatwave, a precise correlation with neonatal health outcomes has not yet been thoroughly investigated.^32^ The scarcity of historical data on neonatal health further complicates the exploration of such associations. By utilizing newly digitized historical weather data and neonatal health records from Lausanne in 1911, this study will contribute to closing a critical research gap in the field of environmental epidemiology. The combination of detailed daily temperature records with neonatal health data offers a unique opportunity to examine the effects of extreme weather events on neonatal outcomes in a historical context, providing valuable insights into the long-term impact of climate change on maternal and neonatal health.

Our objective is to assess the association between prenatal temperature exposure and infant health in Lausanne around 1911. We investigate whether birth weight and the duration of gestation vary depending on maternal exposure to low and high temperature. We additionally report on birth weight and gestational age as binary outcomes: low birth weight (LBW, <2,500g) and preterm birth (PTB, <37 weeks), as well as on continuous placenta weight, birth length and head circumference. Based on the literature, we hypothesize that both low and high temperature had detrimental effects on infant health, potentially lowering infant size and increasing the risks of preterm birth and low birth weight. In addition, we expect that exposure during the last trimester will have the most important effect on infant health. Rates of stillbirth and early neonatal mortality are also described.

## 2. Materials and Methods

### Historical context: Switzerland and Lausanne around 1910

Switzerland had already established itself as one of the wealthiest nations in Europe in terms of Gross Domestic Product (GDP) by the early 20th century.^33^ The period since the late 19th century has seen a rise in per capita income and a significant increase in urbanization. This article focuses on a large city in southwestern of Switzerland, Lausanne. Between 1890 and 1925, the population of Lausanne, a predominantly French-speaking city, doubled from about 34,000 to about 70,000. During this period, several indicators of the standard of living showed positive trends. Mortality rates, including infant and child mortality, declined.^34^ In addition, life expectancy increased significantly, with a child born in the 1920s expected to live approximately 59.1 years^35^ and infectious diseases declined as part of the epidemiological transition and marked improvements in the area of sanitation and hygiene.^35–38^

### Daily temperatures in Lausanne 1908-1913

For daily temperature measurements in Lausanne, we rely on the official cantonal measuring station Champs-de-l’Air (Institut Agricole) in Lausanne. Their measurement data is published in reports for earlier years up to 1908 ^39^, but for the period around 1910, we had to transcribe daily weather bulletins from the station from daily newspapers. Due to occasional poor readability and certain gaps, we used the identical weather bulletins in two daily newspapers for this purpose, the *Nouvelliste vaudois* (taken from https://scriptorium.bcu-lausanne.ch) and *Gazette de Lausanne* (taken from https://www.letempsarchives.ch) to transcribe daily minimum and maximum temperatures between January 1^st^, 1908 and October 31^st^, 1913. Daily mean temperature was calculated by averaging minimum and maximum temperatures. While in the years 1908 to 1913 the maximum temperatures only exceeded 30 degrees Celsius on a few days, the summer of 1911 stands out (Figure 1). For instance, in the months of July to September 1909 and 1910, the thermometer at the measuring station did not exceed 30°C on any day and only exceeded 27.5°C on six days, in the summer of 1911, there were 25 days over 30°C and 52 days over 27.5°C over the same period (Figure 1).

**Figure 1:**
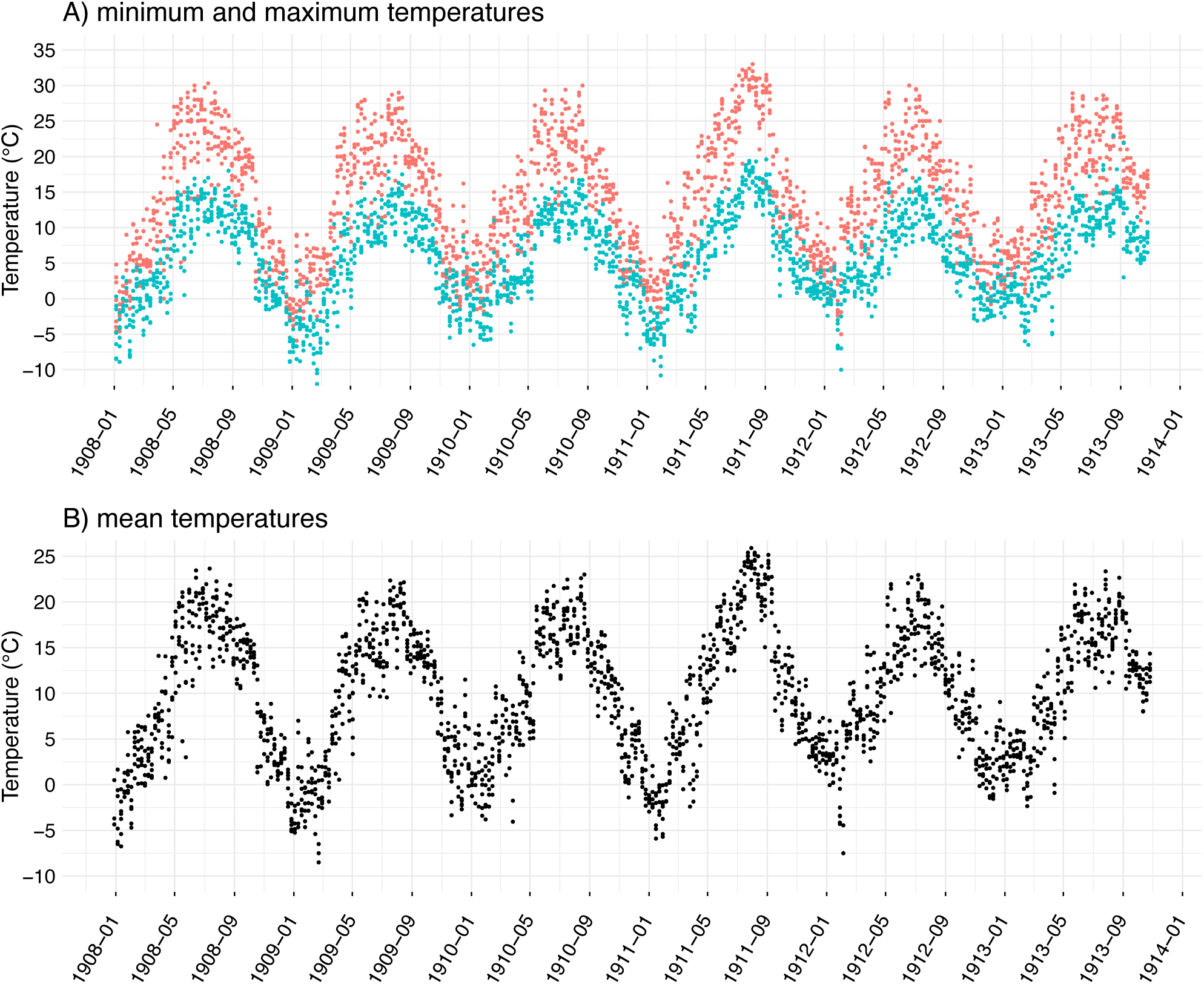
Daily minimum, maximum and mean temperature in Lausanne, January 1^st^, 1908 - October 31^st^ 1913.

### Maternity hospital sources

This article takes advantage of the meticulous documentation and preservation of birth records from Lausanne maternity hospital during the early 20th century ^40,41^. This hospital was the only maternity hospitals in its canton, and it was catered to women from different socio-economic classes, including those who had complicated births. Due to the increasing number and share of hospital births at that time, the maternity hospital had to be expanded between 1900 and 1920. The Lausanne Hospital became a university hospital in 1890, and by 1920 it was responsible for 66% of the city’s births.^42–45^ This publication is part of a larger project during which the birth books of the Lausanne maternity hospital are being transcribed. For this publication the entries of the years 1908-1913 were included, contained in the books KVIII e 197-214 ^46^. Not transcribed were non-birth-related cases, abortions, twin births, and few incomplete records. These maternity books were previously used in studies to research other questions, such as the effect of infectious diseases on infant health ^40,41^.

### Variables

The following variables were transcribed: date of birth, date of last menstrual period, maternal age (years), height (cm), waist circumference (cm), birth weight (grams), placenta weight (grams), gestational age (GA, in weeks) assessed at birth, neonatal sex, maternal gravidity (categorized into 1, 2, >2), civil status (categorized into “married” or “single”, the latter grouping together “single”, “widowed” or “divorced” mothers), stillbirth (corresponds to the living status of the neonate right after delivery) and neonatal mortality. The source contains information about the neonate and its mother for the duration of the mother’s stay in hospital, which varied across mother–infant pairs. Information on early neonatal mortality (nowadays defined as mortality in the first 7 days of life) is missing for pairs who left the maternity before 7 days post-partum. Thus, we use information concerning the first 5 days after delivery only, since virtually all mother-infant pairs stayed for that duration. Maternal gravidity count included previous stillbirths and miscarriages. When the GA calculated based on date of last menstruation and GA assessed at birth matched closely, we kept the derived GA only. However, in case of discrepancy between the two values, we used GA assessed at birth. From this derived GA variable, we defined a PTB variable (GA <37 weeks). Maternal address at the time of delivery was transcribed and a variable “living inside Lausanne” was defined, taking the values “yes”, “no”, or “unsure”.

Some qualitative information on the mother’s general health was recorded. Based on this, we derived the following binary variables: goitre, rickets, infection during pregnancy (defined as any mention of: flu, syphilis, gonorrhoea or tuberculosis). In addition, a categorical variable was created to describe maternal morphology (“thin”, “obese” or “neither”). Maternal occupation was collected and coded using the Historical International Standard of Classification of Occupations (HISCO) database ^47,48^. HISCO codes were then grouped into 3 classes: 1 for non-manual occupations, higher managers/professionals, 2 for medium-skilled workers, farmers, and 3 for unskilled workers/farm workers.

### Outcome variables

The main outcomes were birth weight and gestational age as continuous outcomes. Other outcomes are reported on in the supplementary material: PTB, LBW, as binary variables, and placenta weight, birth length and head circumference as continuous variables. Rates of stillbirth and neonatal mortality are only reported descriptively.

### Exclusions

Between January 1^st^, 1908 and December 31^st^, 1913, the database included 2,372 births. From these, we excluded births that took place at home (n=10) and infants with birth weight <500g or gestational age <22 weeks (*n*=4), since these are classified as miscarriages and not stillbirths. Only births for which full pregnancy temperature exposure was available were included (i.e., birthdate before or on October 31^st^, 1913, n= 358 births excluded). The final dataset for our study population consists of 2,000 births.

### Statistical methods

#### Exposure

Mean daily temperature was aggregated at the weekly level, and for each pregnancy it was averaged over the whole pregnancy or during each trimester (1^st^: week 1-13, 2^nd^: week 14-27, 3^rd^: week 28 until delivery). The distribution of temperature over the whole pregnancy has less variation than over each trimester (see percentiles and corresponding temperatures in Table 2); temperature distribution was similar in each trimester. Temperature was modelled with a natural spline using three knots at the 10^th^, 50^th^ and 90^th^ percentile of temperature distribution of whole pregnancies or of the first trimester, to assess the effects of extreme temperatures and consider the non-linearity between temperature exposure and health outcomes. In following models, only livebirths were considered.

#### Continuous outcomes

The effect of mean temperature during the whole pregnancy or during each trimester was assessed using multivariable generalized linear models (GLM). *Binary outcomes*: Cox proportional hazard regression models were used to measure the association between temperature exposure during the whole pregnancy, and the risks of PTB or LBW. Gestational duration was used as the time scale. PTB and LBW were treated as time-to-event outcomes, with term births censored at 37 weeks for the PTB models. The exposure during each trimester was not considered due to the small number of events.

### Adjustment for covariates

Continuous birth weight outcome model was first adjusted for a set of variables that were expected to be associated with it: season of conception, maternal age, height, civil status and gravidity, and infant sex, based on the literature. There is an important seasonal variation in neonatal health, including mean birth weight ^49^; it is preferable to control for season of conception rather than season of birth ^50^. Another set of potential explanatory and/or confounding variables (maternal morphology, rickets, goitre, HISCO class and Lausanne residency status) were added using a stepwise algorithm. Based on the lowest Akaike information criterion (AIC), the model was adjusted for season of conception, maternal age, height, civil status, Lausanne residency status, morphology and gravidity, and infant sex. All other models (with outcomes gestational age, placenta weight, birth length, head circumference, PTB, LBW) were adjusted for the same variables than the birth weight outcome model. We included all trimesters in a single model rather than using separate models for each trimester (for which there are biased results due to the seasonality of exposure and the correlation between the time windows – i.e. the trimesters) ^51^.

#### Sensitivity analyses

For the main outcomes (continuous birth weight and gestational age), temperature exposure during the whole pregnancy was also modelled using either a cubic-spline or a natural spline, and using different knot combinations (at the tertiles, at the quartiles, or at the 5^th^, 50^th^ and 95^th^ percentiles). Results were similar; although model fit was slightly better (lower AIC) for the models with three knots at the 5^th^, 50^th^ and 95^th^ percentiles, there were too few births exposed to these extremes. Therefore, we chose to use the 10^th^, 50^th^ and 90^th^ percentiles knots.

## 3. Results

The descriptive statistics for the 2,000 mother-child pairs from the Lausanne Maternity Hospital between 1909 and 1913 are presented in **Table 1**. The mothers were on average 28.1 years (SD=6.7) old and 155.6 cm (SD=6.6 cm) tall. More than 70% of them were married. Overall, 34% of the deliveries were first births and 45% of the births were third or subsequent births. Most mothers lived inside the city of Lausanne and belonged to the middle HISCO class. The infants weighed 3076 grams on average (SD=600), and the mean placenta weight was 560 grams (SD=125). Over the entire observation period, the LBW rate was 13%, the PTB was 12%, 5% of the births were stillbirths, and around 4% of the newborns died in the hospital in the first five days.

**Table 1:** Maternal and neonatal characteristics. ^1^ Mean (SD); n (%)

| Maternal variables | *n* = 2, 000<sup>1</sup> | Neonatal variables | *n* = 2,000<sup>1</sup> |
| --- | --- | --- | --- |
| maternal age (years) | 28.1 (6.7) | birth weight (g) | 3,075.7 (600.0) |
| height (cm) | 155.6 (6.6) | head circumference (cm) | 34.1 (2.5) |
| missing | 79 | missing | 40 |
| civil status |  | placenta weight | 559.6 (124.8) |
| married | 1,469 (73%) | missing | 13 |
| single or missing | 531 (27%) | birth length (cm) | 48.8 (3.1) |
| parity |  | missing | 7 |
| 1 | 682 (34%) | ponderal index | 2.6 (1.1) |
| 2 | 420 (21%) | missing | 7 |
| >2 | 898 (45%) | gestational age (weeks) | 38.9 (2.4) |
| living in Lausanne |  | sex |  |
| yes | 1,043 (52%) | male | 1,026 (51%) |
| no or unsure | 957 (48%) | female | 972 (49%) |
| goitre | 149 (7%) | missing | 2 |
| rickets | 172 (9%) | low birth weight (<2'500g) | 257 (13%) |
| infection | 86 (4%) | preterm birth (<37 weeks) | 239 (12%) |
| HISCO class |  | stillbirth | 100 (5%) |
| 1 | 346 (17%) | neonatal mortality d1-5 | 85 (4%) |
| 2 | 1,529 (76%) | missing | 5 |
| 3 | 37 (2%) |  |  |
| missing | 88 (4%) |  |  |
| season of conception |  |  |  |
| spring | 456 (23%) |  |  |
| summer | 529 (26%) |  |  |
| autumn | 521 (26%) |  |  |
| winter | 494 (25%) |  |  |

**Table 2:** Neonatal characteristics depending on percentiles of temperature exposure during the whole pregnancy and during each trimester. For continuous outcomes, the mean is displayed; for categorical outcomes, the number and the percentage are displayed. Note: for the third trimester, the number of exposed infants does not sum up to n=2,000, because 5 pregnancies ended before the third trimester (<26 weeks).

|  | Whole pregnancy temperature |  |  | First trimester |  |  | Second trimester |  |  | Third trimester |  |  |
| --- | --- | --- | --- | --- | --- | --- | --- | --- | --- | --- | --- | --- |
| percentile | <10th | >90th | 10-90th | <10<sup>th</sup> | >90<sup>th</sup> | 10-90th | <10th | >90th | 10-90th | <10th | >90th | 10-90th |
| Corresponding temperature (°C) | 10.5 | 16.8 | 10.5-16.8 | 4.7 | 22.4 | 4.7-22.4 | 5.1 | 21.9 | 5.1-21.9 | 5.4 | 21.8 | 5.4-21.8 |
| n | 203 | 208 | 1,589 | 198 | 196 | 1,606 | 193 | 197 | 1,610 | 200 | 202 | 1,593 |
| birth weight (g) | 2,870.3<br>(630.9) | 2,784.5<br>(735.4) | 3,140.1<br>(558.1) | 3,124.2<br>(598.6) | 3,091.2<br>(591.5) | 3,067.9<br>(601.3) | 3,112.4<br>(565.2) | 3,058.7<br>(568.3) | 3,073.4<br>(608.0) | 3,010.8<br>(680.0) | 2,903.5<br>(638.7) | 3,111.8<br>(570.0) |
| head circumference (cm) | 34.0<br>(2.5) | 33.3<br>(2.6) | 34.3<br>(2.4) | 34.5<br>(2.0) | 34.4<br>(2.1) | 34.1<br>(2.6) | 34.7<br>(2.0) | 34.2<br>(1.9) | 34.1<br>(2.6) | 34.3<br>(3.1) | 33.7<br>(2.3) | 34.2<br>(2.3) |
| placenta weight | 543.2<br>(130.1) | 513.8<br>(116.0) | 567.6<br>(123.8) | 579.4<br>(127.2) | 555.8<br>(119.1) | 557.6<br>(125.0) | 580.6<br>(131.3) | 542.4<br>(116.7) | 559.1<br>(124.7) | 559.8<br>(126.7) | 548.4<br>(138.4) | 561.4<br>(122.5) |
| birth length (cm) | 47.9<br>(3.5) | 47.2<br>(4.3) | 49.2<br>(2.8) | 49.1<br>(2.7) | 48.8<br>(2.8) | 48.8<br>(3.2) | 49.2<br>(2.8) | 48.7<br>(3.3) | 48.8<br>(3.1) | 48.6<br>(3.6) | 48.2<br>(3.3) | 49.0<br>(2.9) |
| gestational age (weeks) | 38.0<br>(2.7) | 37.3<br>(3.7) | 39.2<br>(2.1) | 39.0<br>(2.1) | 39.0<br>(2.4) | 38.8<br>(2.5) | 38.9<br>(2.0) | 38.9<br>(2.4) | 38.8<br>(2.5) | 38.4<br>(3.0) | 38.2<br>(2.8) | 39.0<br>(2.2) |
| sex |  |  |  |  |  |  |  |  |  |  |  |  |
| male | 100<br>(50%) | 103<br>(50%) | 823<br>(52%) | 93 (47%) | 101<br>(52%) | 832<br>(52%) | 102<br>(53%) | 106<br>(54%) | 818<br>(51%) | 99 (50%) | 90 (45%) | 834<br>(52%) |
| female | 102<br>(50%) | 105<br>(50%) | 765<br>(48%) | 105<br>(53%) | 95 (48%) | 772<br>(48%) | 91 (47%) | 91 (46%) | 790<br>(49%) | 100<br>(50%) | 112<br>(55%) | 758<br>(48%) |
| low birth weight (<2'500g) | 49 (24%) | 53 (25%) | 155<br>(10%) | 25 (13%) | 22 (11%) | 210<br>(13%) | 26 (13%) | 21 (11%) | 210<br>(13%) | 30 (15%) | 44 (22%) | 178<br>(11%) |
| preterm birth (<37 weeks) | 46 (23%) | 54 (26%) | 139 (9%) | 20 (10%) | 23 (12%) | 196<br>(12%) | 24 (12%) | 21 (11%) | 194<br>(12%) | 30 (15%) | 37 (18%) | 167<br>(10%) |
| stillbirth | 13 (6%) | 19 (9%) | 68 (4%) | 11 (6%) | 11 (6%) | 78 (5%) | 7 (4%) | 11 (6%) | 82 (5%) | 16 (8%) | 13 (6%) | 68 (4%) |
| early neonatal mortality d1-5 | 12 (6%) | 15 (7%) | 58 (4%) | 10 (5%) | 8 (4%) | 67 (4%) | 10 (5%) | 5 (3%) | 70 (4%) | 10 (5%) | 16 (8%) | 57 (4%) |

<sup>1</sup>Mean (SD); n (%)

In terms of how many infants have been exposed to <10^th^ and/or >90^th^ percentile of temperature across the individual years of observation, **Table S2** shows that for each birthyear of the dataset (1908-1913), a total of n=208 infants were exposed to the highest daily mean temperatures (>90^th^ percentile, 16.8°C) during the whole pregnancy. Most infants exposed to these high temperatures were born in 1911 (43%) and 1912 (25%), i.e. they were exposed to the extreme heatwave in July to September 1911. On the other hand, infants born in 1908 or 1912 were not exposed to the coldest temperatures (<10th percentile, <10.5°C) during the entire gestation. When comparing neonatal health outcomes of these infants exposed to temperature percentiles <10^th^ or >90^th^ with those of infants exposed to temperature percentiles 10–90^th^, we see consistently worse outcomes for both exposure during the entire pregnancy and the third trimester (**Table 2**). This concerns birth weight, head circumference, placenta weight, as well as gestational age (please note that this is not adjusted for any covariates). LBW (24% for <10^th^ percentile of temperature exposure and 25% for >90^th^ percentile of temperature exposure compared to 10% for 10–90^th^ percentile), PTB (23% and 26% compared to 9%) and stillbirth rate (6% and 9% compared to 4%) were also higher when exposed to temperature percentiles <10^th^ or >90^th^. **Table S1** shows that maternal factors such as age, height, civil status, HISCO class etc. do not differ notably between temperature exposure groups.

The predicted values of the continuous outcome variables birth weight and gestational age depending on temperature exposure during the whole pregnancy and during each trimester from generalized linear models are reported in **Table 3** and the effects are visualized in **Figure 2**. The negative and n-shaped effect of heat and cold temperature exposure is particularly strong for the whole pregnancy and the third trimester exposures. Infants exposed to the 95^th^ percentile of temperature during the whole pregnancy were −211.3 grams lighter (95%CI −284.8 to −137.9) and were born −1.1 weeks (95%CI −1.3 to −0.8) earlier than infants who were exposed to median temperature during the whole pregnancy. The effect is much less pronounced for cold weather than for hot temperature exposure and does not appear when exposure is considered only in the second trimester. The results for placenta weight, birth length, and head circumference are shown in **Table S3** and **Figure S1** and follow a similar pattern.

**Figure 2:**
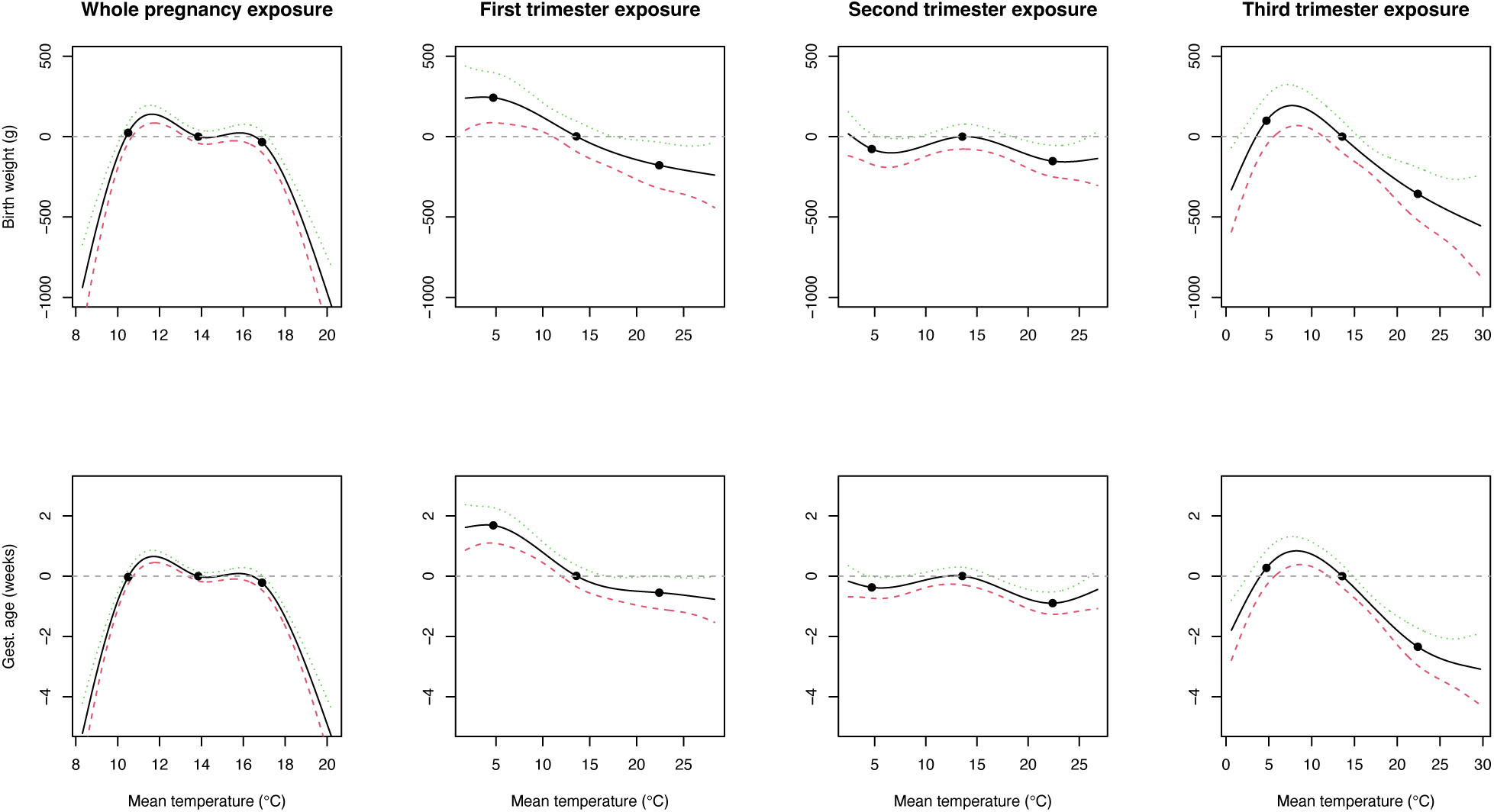
Effect of temperature variation on birth weight (top) and gestational age (bottom), based on generalized linear models. For each outcome, there is one model considering whole pregnancy exposure (first column) and one model considering trimester exposure (three other columns) with trimesters in the same model. Black dots: 10^th^, 50^th^ and 90^th^ percentiles of the whole pregnancy temperature or trimester temperature distribution (correspond to the first trimester distribution); dashed green and red lines: 95% confidence intervals; dashed grey line: y=0, correspond to no variation of the outcomes. Note: only livebirths are considered.

**Table 3:** Predicted values of birth weight and gestational age depending on temperature exposure during the whole pregnancy and during each trimester, from generalized linear models (corresponding to Figure 2). The references of covariates were the median for continuous variables, and the most common category for the categorical variables (maternal age: 27 years old, maternal height: 156cm, gravidity: >2, conception season: autumn, morphology: neither thin nor obese, civil status: married, residency: living in Lausanne, sex: male).

| Window of exposure | Whole pregnancy exposure model |  |  |  |  | Trimester-exposure model |  |  |  |  |  |  |  |  |  |
| --- | --- | --- | --- | --- | --- | --- | --- | --- | --- | --- | --- | --- | --- | --- | --- |
|  | Whole pregnancy |  |  |  |  | First trimester |  |  |  | Second trimester |  |  | Third trimester |  |  |
| Birth weight (g) | Perc. | T°C | beta | lci | uci | T°C | beta | lci | uci | beta | lci | uci | beta | lci | uci |
|  | 1st | 9.48 | -330.02 | -443.21 | -216.82 | 2.06 | 221.30 | 30.59 | 412.01 | 112.57 | -36.77 | 261.91 | -85.17 | -287.83 | 117.49 |
|  | 5th | 10.20 | -58.16 | -124.99 | 8.66 | 3.52 | 226.11 | 62.04 | 390.17 | 44.13 | -63.46 | 151.73 | 71.53 | -94.49 | 237.55 |
|  | 10th | 10.51 | 24.08 | -37.75 | 85.91 | 4.72 | 222.63 | 66.76 | 378.51 | 0.54 | -94.65 | 95.72 | 169.97 | 17.02 | 322.93 |
|  | 50th | 13.85 | -2.95 | -44.71 | 38.80 | 13.57 | -17.81 | -112.33 | 76.71 | 77.45 | 0.13 | 154.78 | 71.14 | -27.45 | 169.73 |
|  | 90th | 16.82 | -28.47 | -93.67 | 36.73 | 22.40 | -197.68 | -341.31 | -54.04 | -74.79 | -172.56 | 22.98 | -285.32 | -449.32 | -121.32 |
|  | 95th | 17.79 | -211.31 | -284.76 | -137.86 | 24.98 | -226.96 | -377.77 | -76.14 | -74.46 | -195.95 | 47.03 | -362.21 | -547.92 | -176.51 |
|  | 99th | 18.85 | -533.77 | -656.34 | -411.20 | 27.39 | -250.28 | -431.61 | -68.95 | -52.62 | -239.99 | 134.75 | -425.74 | -655.81 | -195.67 |
| Gestational age (weeks) | 1st | 9.48 | -1.90 | -2.32 | -1.47 | 2.06 | 1.27 | 0.55 | 1.99 | 0.28 | -0.29 | 0.84 | -0.36 | -1.13 | 0.40 |
|  | 5th | 10.20 | -0.42 | -0.68 | -0.17 | 3.52 | 1.33 | 0.71 | 1.95 | 0.12 | -0.28 | 0.53 | 0.39 | -0.24 | 1.02 |
|  | 10th | 10.51 | 0.03 | -0.20 | 0.26 | 4.72 | 1.32 | 0.73 | 1.91 | 0.04 | -0.32 | 0.40 | 0.88 | 0.30 | 1.45 |
|  | 50th | 13.85 | 0.04 | -0.12 | 0.20 | 13.57 | -0.36 | -0.71 | 0.00 | 0.41 | 0.12 | 0.70 | 0.60 | 0.23 | 0.98 |
|  | 90th | 16.82 | -0.13 | -0.38 | 0.11 | 22.40 | -0.91 | -1.46 | -0.37 | -0.49 | -0.86 | -0.12 | -1.74 | -2.36 | -1.12 |
|  | 95th | 17.79 | -1.06 | -1.34 | -0.78 | 24.98 | -0.99 | -1.56 | -0.42 | -0.29 | -0.75 | 0.17 | -2.12 | -2.82 | -1.42 |
|  | 99th | 18.85 | -2.67 | -3.14 | -2.21 | 27.39 | -1.09 | -1.78 | -0.41 | 0.06 | -0.65 | 0.76 | -2.34 | -3.21 | -1.47 |

Consistently with the continuous outcome models, PTB and LBW hazard ratios (HR) increase at high temperatures (**Figure S2**), but confidence intervals are wide, and the HR are implausibly high at the most extreme exposures (HR >10). The instant risk of birth at week X of gestation (X<37 weeks) is multiplied by 2.56 [95%CI 1.53 to 4.27] for pregnancies exposed to the 1^st^ percentile of highest temperatures (>18.85°C, **Table S4**). The instant risk of birth with a weight <2,500g is multiplied by 1.824 [95%CI 1.15 to 2.89] for pregnancies exposed to the 5^th^ percentile of highest temperatures (17.79°C). On the other hand, exposure to the lowest temperature (<1^st^ percentile, i.e. 9.48°C) is associated with a higher risk of birth with a LBW (HR 4.98 [95%C 3.11 to 7.99] but not with birth before the term (HR 0.95 [95%CI 0.62 to 1.45]) (**Figure S2**, **Table S4**).

## 4. Discussion

In this historical data set of 2,000 mother–infant pairs from Lausanne (1909–1913), exposure to extreme temperatures during pregnancy - particularly in the third trimester - was associated with consistently poorer birth outcomes. Compared with infants exposed to moderate temperatures (10–90th percentile), those exposed to heat (>90th percentile) or cold (<10th percentile) throughout gestation had lower birth weight, shorter gestation, smaller head circumference, and reduced placenta weight, alongside higher rates of low birth weight, preterm birth, and stillbirth. The strongest effects were observed for heat exposure (and particularly during the 1911 extreme heat wave), with whole-pregnancy exposure to the 95th percentile linked to a marked mean birth weight and gestation reduction. Hazard ratios for LBW and PTB generally increased with high temperatures, although confidence intervals widened at the extremes.

Only a limited number of studies using historical datasets from the early 20th century and Central Europe are available to compare with our results. However, our results are partially in line with a study analysing 14,000 deliveries and their association with daily ambient temperature in Uppsala (Sweden) during the years 1915 to 1929: The authors found that the risk for stillbirth and PTB increased and birth length was lower when ambient temperature fell, but no association was found with birth weight and gestational age, while warm temperature extremes were associated with higher risk of PTB ^52^. Such differences need to be better understood in comparative studies, but it could be that the latitude and thus the completely different temperature environment plays a role here. Our findings add to the literature that not only warmth but also heat, in addition to cold, was associated with negative health consequences for infants in a historical setting.

These associations between both heat and cold and neonatal health have been demonstrated on numerous occasions in modern data sets, as has the fact that timing, and particularly the third trimester, appears to play an important role and that there is a dose-response relationship (the hotter and longer the exposure, the more negative the effect) ^6–8,12–14^. The mechanisms behind these associations are likely to be manifold: Pregnant women may be more susceptible to extreme temperatures because of inefficient thermoregulation due to weight gain, decreased surface area to mass ratio, and a higher metabolic heat production ^53^; in addition, cytokines which are involved in labour induction may be released due to heat. As a response to heat stress, the blood is redistributed to the skin to dissipate the heat, which may in turn impair oxygen supply to the foetus may be impaired ^54^. Also, it has been postulated that heat exposure may be associated with PTB due to channels such as hydration, hormone secretion, altered blood viscosity, premature rupture of membranes, or reduced thermoregulation of the pregnant women ^55^. Meanwhile, exposure to cold temperatures may restrict blood flow to the placenta and thus impar foetal growth ^53,56^.

Our study has several limitations that should be taken into account when interpreting the results. Firstly, before World War I, the proportion of children born in hospitals was still relatively low (estimates are around 34%); it was only during and after World War I that this proportion rose to around 66%. On the one hand, this maternity hospital was the only one in the city of Lausanne, but on the other hand, we are missing all the births that took place at home. We also cannot accurately assess whether there was a socio-economic selection bias and whether certain subgroups of women gave birth in hospital more or less frequently. However, we know with certainty that there were never any restrictions on admission to the hospital for childbirth and that both problematic and uncomplicated births took place there. Because the lower coverage of the source also affected the absolute number of births, we focused on weekly births. Secondly, our temperature data is limited to only one measuring station in Lausanne. We thus have to assume an equal temperature exposure for all pregnant women of Lausanne and its surroundings, however temperature exposure may actually have varied depending on housing conditions, proximity to the lake of Geneva or to vegetation. Furthermore, almost half of the women were living outside of the city of Lausanne and could have been exposed to even more different temperatures in rural or mountainous areas, spared from the urban heat island effect. Even while this limitation should bias associations towards the null, especially in the case of a rather small sample size, we still find evidence that extreme temperature exposure during pregnancy was associated with adverse neonatal health.

## Conclusion

Current vulnerability research in the context of extreme temperatures tends to focus on the elderly population. However, there is growing evidence that temperature exposure during pregnancy has negative effects on newborn health, which means that the youngest age groups should also be included in vulnerability considerations ^57^. Here, we use a historical example from a completely different life context from over 100 years ago to show that this could be a constant over time. Further similar studies on historical birth data would need to follow in order to verify the generalizability of our results.

## Acknowledgements

The authors would like to thank Urmila Bhattacharyya, Joel Floris, Antonia Galliker, Nora Haag, and Rosa Hinselmann for participating to the data transcription; the Cantonal archives of Vaud and its archivists for the access to the original data; Frank Rühli, Sabine Rohrmann and Viktor von Wyl for their helpful comments; Nathalie Costet and Valérie Gares for helpful Cox models discussions.

## Author statements

### Data Availability Statement

The data that support the findings of this study are available from the corresponding author upon reasonable request. The codes used for the data analysis are publicly available in the following online repository (Github): Link follows.

### Ethics committee approval

Ethics approval was obtained from the Zurich Ethic Commission (BASEC Number 2021-00628).

## Supplementary Material

**Figure S1:**
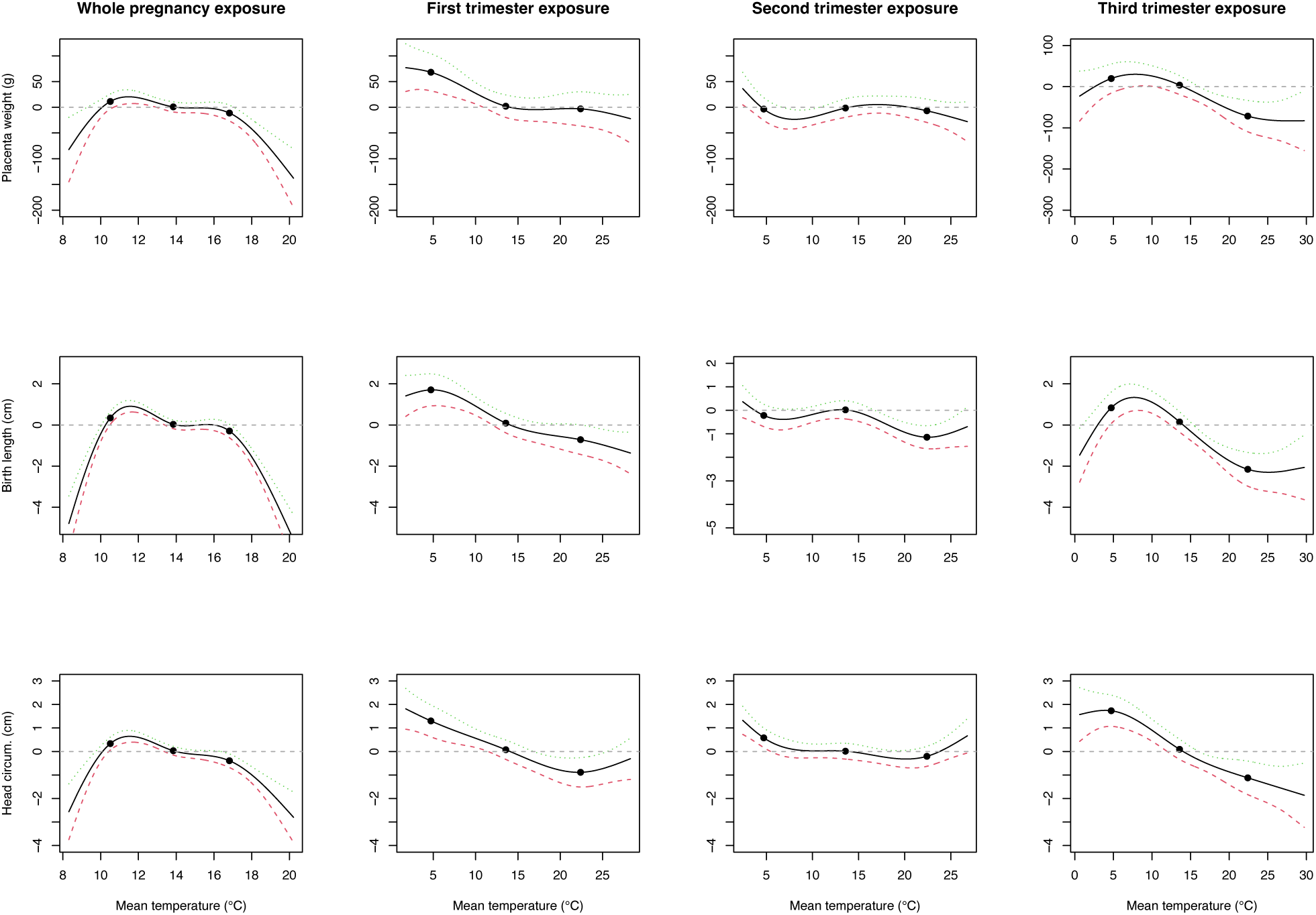
Effect of temperature variation on placenta weight (top), birth length (middle) and head circumference (bottom), based on generalized linear models. For each outcome, there is one model considering whole pregnancy exposure (first column) and one model considering trimester exposure (three other columns) with trimesters in the same model. Black dots: 10^th^, 50^th^ and 90^th^ percentiles of the whole pregnancy temperature or trimester temperature distribution (correspond to the first trimester distribution); dashed green and red lines: 95% confidence intervals; dashed grey line: y=0, correspond to no variation of the outcomes. Note: only livebirths are considered.

**Figure S2:**
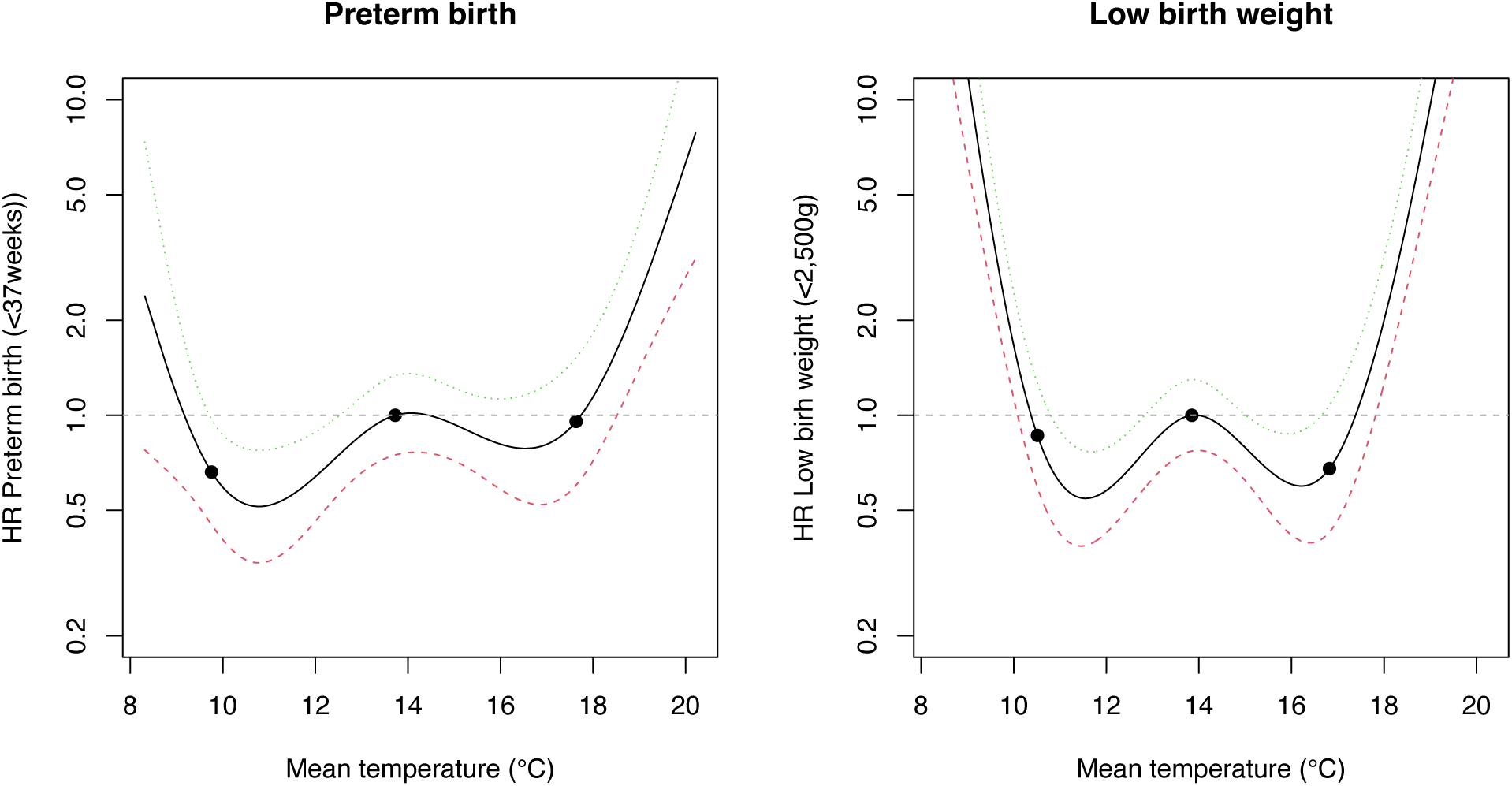
Effect of temperature variation on the instant risk of birth before 37 weeks of gestation (left) or with a low birth weight (<2,500g, right), based on Cox proportional hazard ratio models. For each outcome, there is one model considering whole pregnancy exposure. Black dots: 10^th^, 50^th^ and 90^th^ percentiles of the whole pregnancy temperature; dashed green and red lines: 95% confidence intervals; dashed grey line: y=0, correspond to no variation of the outcomes. Note: only livebirths are considered.

**Table S1:**
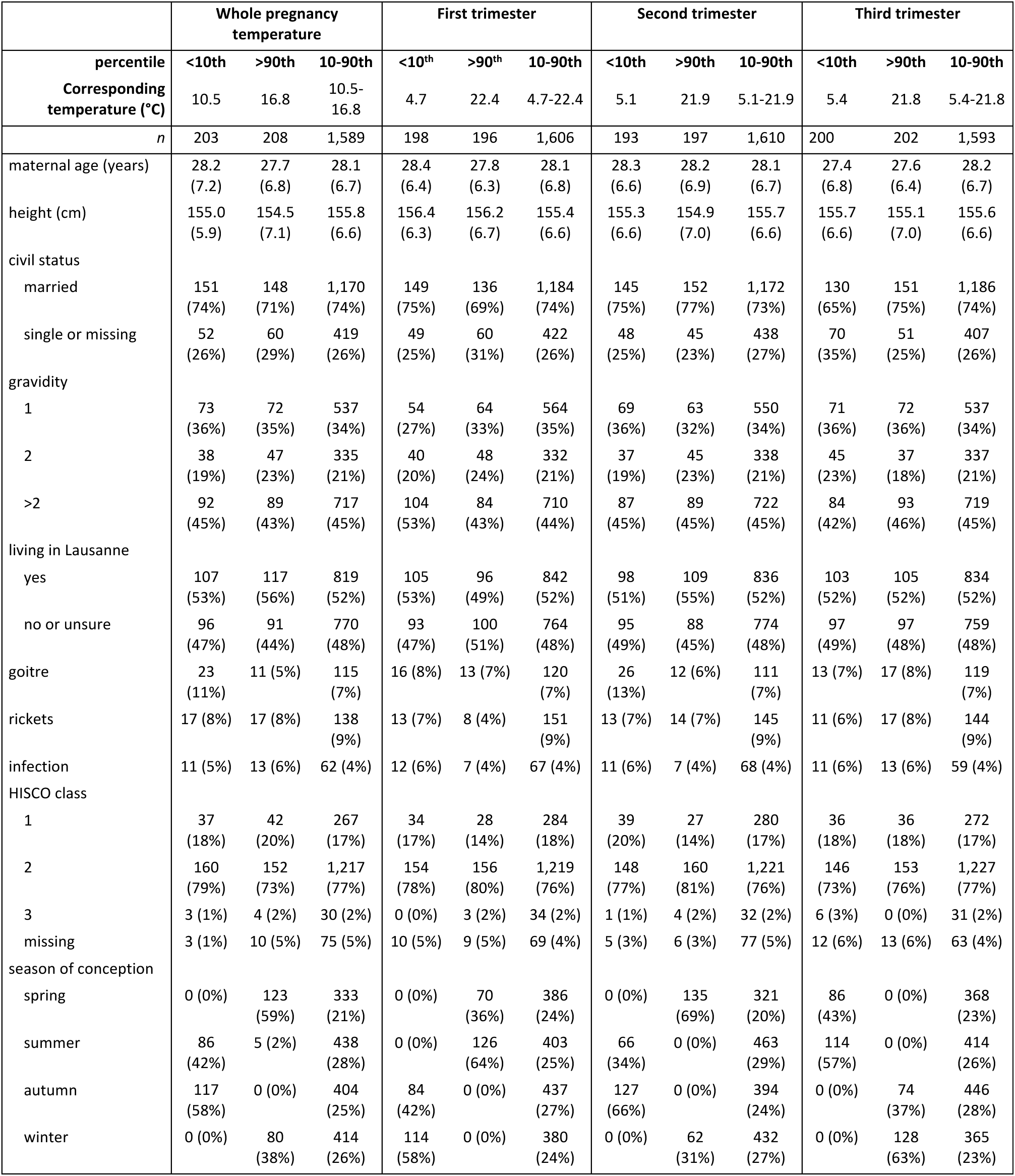
maternal characteristics depending on percentiles of temperature exposure during the whole pregnancy and during each trimester. For continuous outcomes, the mean is displayed; for categorical outcomes, the number and the percentage are displayed. Note: for the third trimester, the number of exposed mothers does not sum up to n=2,000, because 5 pregnancies ended before the third trimester (<26 weeks).

**Table S2:** Number and frequencies of infants that have been exposed to each percentile of temperature during the whole pregnancy or during each trimester, depending on the year of birth. Perc.: percentile.

| Birthyear | Perc. of temperature exposure | Whole pregnancy |  | Trimester 1 |  | Trimester 2 |  | Trimester 3 |  |
| --- | --- | --- | --- | --- | --- | --- | --- | --- | --- |
|  |  | Count | Frequency | Count | Frequency | Count | Frequency | Count | Frequency |
| 1908 | <10th | 0 | 0 | 11 | 5,56 | 0 | 0 | 0 | 0 |
| 1909 | <10th | 105 | 51,72 | 91 | 45,96 | 101 | 52,33 | 87 | 43,5 |
| 1910 | <10th | 17 | 8,37 | 28 | 14,14 | 18 | 9,33 | 34 | 17 |
| 1911 | <10th | 25 | 12,32 | 59 | 29,8 | 64 | 33,16 | 62 | 31 |
| 1912 | <10th | 0 | 0 | 0 | 0 | 0 | 0 | 3 | 1,5 |
| 1913 | <10th | 56 | 27,59 | 9 | 4,55 | 10 | 5,18 | 14 | 7 |
|  | sum | 203 | 100% | 198 | 100% | 193 | 100% | 200 | sum: 100% |
| 1908 | >90th | 28 | 13,46 | 0 | 0 | 47 | 23,86 | 12 | 5,94 |
| 1909 | >90th | 15 | 7,21 | 74 | 37,76 | 23 | 11,68 | 19 | 9,41 |
| 1910 | >90th | 14 | 6,73 | 0 | 0 | 19 | 9,64 | 31 | 15,35 |
| 1911 | >90th | 89 | 42,79 | 3 | 1,53 | 61 | 30,96 | 92 | 45,54 |
| 1912 | >90th | 51 | 24,52 | 119 | 60,71 | 46 | 23,35 | 18 | 8,91 |
| 1913 | >90th | 11 | 5,29 | 0 | 0 | 1 | 0,51 | 30 | 14,85 |
|  | sum | 208 | 100% | 196 | 100% | 197 | 100% | 202 | 100% |
| 1908 | 10-90th | 52 | 3,27 | 69 | 4,3 | 33 | 2,05 | 68 | 4,27 |
| 1909 | 10-90th | 263 | 16,55 | 218 | 13,57 | 259 | 16,09 | 277 | 17,39 |
| 1910 | 10-90th | 331 | 20,83 | 334 | 20,8 | 325 | 20,19 | 294 | 18,46 |
| 1911 | 10-90th | 282 | 17,75 | 334 | 20,8 | 271 | 16,83 | 242 | 15,19 |
| 1912 | 10-90th | 379 | 23,85 | 311 | 19,36 | 384 | 23,85 | 408 | 25,61 |
| 1913 | 10-90th | 282 | 17,75 | 340 | 21,17 | 338 | 20,99 | 304 | 19,08 |
|  | sum | 1589 | 100% | 1606 | 100% | 1610 | 100% | 1593 | 100% |

**Table S3:** Predicted values of placenta weight, birth length and head circumference depending on temperature exposure during the whole pregnancy and during each trimester, from generalized linear models (corresponding to Figure S1). The references of covariates were the median for continuous variables, and the most common category for the categorical variables (maternal age: 27 years old, maternal height: 156cm, gravidity: >2, conception season: autumn, morphology: neither thin nor obese, civil status: married, residency: living in Lausanne, sex: male).

| Window of exposure | Whole pregnancy exposure model |  |  |  |  | Trimester-exposure model |  |  |  |  |  |  |  |  |  |
| --- | --- | --- | --- | --- | --- | --- | --- | --- | --- | --- | --- | --- | --- | --- | --- |
|  | Whole pregnancy |  |  |  |  | First trimester |  |  |  | Second trimester |  |  | Third trimester |  |  |
|  | Perc. | T°C | beta | lci | uci | T°C | beta | lci | uci | beta | lci | uci | beta | lci | uci |
| Placenta weight (g) | 1st | 9.48 | -22.96 | -49.53 | 3.60 | 2.06 | 55.31 | 11.01 | 99.60 | 49.87 | 15.32 | 84.42 | 8.68 | -38.16 | 55.53 |
|  | 5th | 10.20 | 3.16 | -12.56 | 18.89 | 3.52 | 51.67 | 13.62 | 89.72 | 21.97 | -2.95 | 46.88 | 24.22 | -14.16 | 62.59 |
|  | 10th | 10.51 | 10.85 | -3.71 | 25.42 | 4.72 | 47.04 | 10.90 | 83.17 | 2.82 | -19.25 | 24.89 | 34.18 | -1.18 | 69.54 |
|  | 50th | 13.85 | 0.33 | -9.51 | 10.18 | 13.57 | -18.98 | -40.86 | 2.90 | 4.83 | -13.14 | 22.81 | 18.24 | -4.64 | 41.13 |
|  | 90th | 16.82 | -11.67 | -27.06 | 3.73 | 22.40 | -24.14 | -57.36 | 9.08 | -0.50 | -23.20 | 22.20 | -57.09 | -95.18 | -19.00 |
|  | 95th | 17.79 | -35.69 | -53.04 | -18.34 | 24.98 | -30.02 | -65.04 | 5.00 | -12.38 | -40.56 | 15.80 | -66.25 | -109.34 | -23.16 |
|  | 99th | 18.85 | -75.69 | -104.62 | -46.76 | 27.39 | -39.33 | -81.77 | 3.11 | -24.77 | -68.14 | 18.60 | -68.72 | -122.00 | -15.45 |
| Birth length (cm) | 1st | 9.48 | -1.58 | -2.15 | -1.01 | 2.06 | 1.11 | 0.15 | 2.06 | 0.93 | 0.18 | 1.68 | -0.20 | -1.22 | 0.81 |
|  | 5th | 10.20 | -0.14 | -0.47 | 0.20 | 3.52 | 1.29 | 0.47 | 2.11 | 0.51 | -0.03 | 1.05 | 0.63 | -0.20 | 1.47 |
|  | 10th | 10.51 | 0.30 | -0.01 | 0.61 | 4.72 | 1.36 | 0.58 | 2.14 | 0.24 | -0.24 | 0.71 | 1.16 | 0.40 | 1.93 |
|  | 50th | 13.85 | -0.01 | -0.22 | 0.20 | 13.57 | -0.26 | -0.73 | 0.21 | 0.47 | 0.09 | 0.86 | 0.48 | -0.02 | 0.97 |
|  | 90th | 16.82 | -0.34 | -0.67 | -0.01 | 22.40 | -1.06 | -1.78 | -0.35 | -0.69 | -1.18 | -0.20 | -1.83 | -2.64 | -1.01 |
|  | 95th | 17.79 | -1.31 | -1.68 | -0.94 | 24.98 | -1.31 | -2.07 | -0.56 | -0.51 | -1.12 | 0.10 | -1.97 | -2.90 | -1.04 |
|  | 99th | 18.85 | -2.98 | -3.59 | -2.36 | 27.39 | -1.60 | -2.51 | -0.69 | -0.14 | -1.08 | 0.79 | -1.90 | -3.05 | -0.75 |
| Head circumference (cm) | 1st | 9.48 | -0.74 | -1.24 | -0.23 | 2.06 | 1.62 | 0.79 | 2.45 | 1.39 | 0.74 | 2.05 | 1.50 | 0.62 | 2.38 |
|  | 5th | 10.20 | 0.08 | -0.22 | 0.37 | 3.52 | 1.36 | 0.64 | 2.07 | 0.88 | 0.41 | 1.35 | 1.57 | 0.85 | 2.29 |
|  | 10th | 10.51 | 0.32 | 0.05 | 0.60 | 4.72 | 1.16 | 0.48 | 1.84 | 0.52 | 0.10 | 0.93 | 1.56 | 0.89 | 2.22 |
|  | 50th | 13.85 | 0.02 | -0.17 | 0.20 | 13.57 | -0.07 | -0.48 | 0.35 | -0.05 | -0.39 | 0.29 | -0.08 | -0.51 | 0.34 |
|  | 90th | 16.82 | -0.41 | -0.70 | -0.12 | 22.40 | -1.03 | -1.65 | -0.40 | -0.27 | -0.69 | 0.16 | -1.30 | -2.01 | -0.58 |
|  | 95th | 17.79 | -0.88 | -1.20 | -0.55 | 24.98 | -0.87 | -1.53 | -0.22 | 0.18 | -0.35 | 0.71 | -1.56 | -2.37 | -0.75 |
|  | 99th | 18.85 | -1.63 | -2.18 | -1.09 | 27.39 | -0.58 | -1.36 | 0.21 | 0.75 | -0.07 | 1.56 | -1.80 | -2.80 | -0.80 |

**Table S4:**
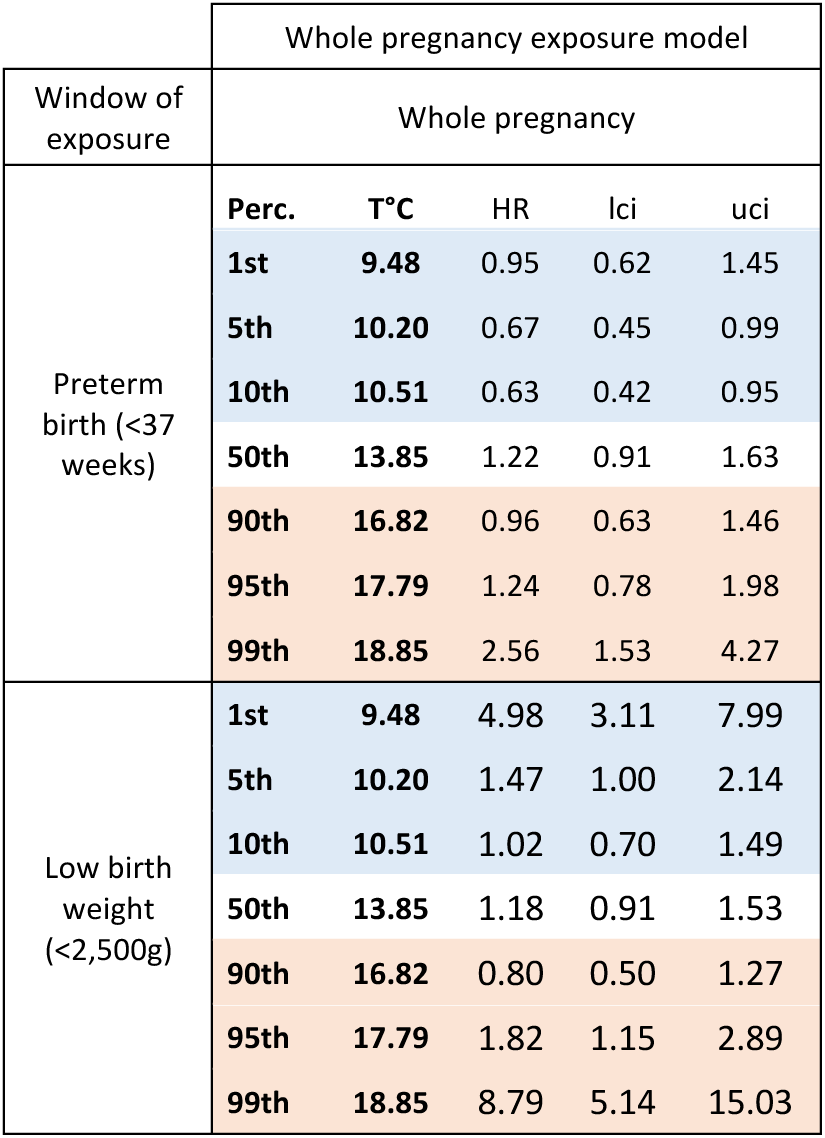
Predicted values of preterm birth and low birth weight risks depending on temperature exposure during the whole pregnancy, from Cox proportional hazard ratio models (corresponding to Figure S2). The references of covariates were the median for continuous variables, and the most common category for the categorical variables (maternal age: 27 years old, maternal height: 156cm, gravidity: >2, conception season: autumn, morphology: neither thin nor obese, civil status: married, residency: living in Lausanne, sex: male).

| Window of exposure | Whole pregnancy exposure model |  |  |  |  |
| --- | --- | --- | --- | --- | --- |
|  | Whole pregnancy |  |  |  |  |
| Preterm birth (<37 weeks) | Perc. | T°C | HR | lci | uci |
|  | 1st | 9.48 | 0.95 | 0.62 | 1.45 |
|  | 5th | 10.20 | 0.67 | 0.45 | 0.99 |
|  | 10th | 10.51 | 0.63 | 0.42 | 0.95 |
|  | 50th | 13.85 | 1.22 | 0.91 | 1.63 |
|  | 90th | 16.82 | 0.96 | 0.63 | 1.46 |
|  | 95th | 17.79 | 1.24 | 0.78 | 1.98 |
|  | 99th | 18.85 | 2.56 | 1.53 | 4.27 |
| Low birth weight (<2,500g) | 1st | 9.48 | 4.98 | 3.11 | 7.99 |
|  | 5th | 10.20 | 1.47 | 1.00 | 2.14 |
|  | 10th | 10.51 | 1.02 | 0.70 | 1.49 |
|  | 50th | 13.85 | 1.18 | 0.91 | 1.53 |
|  | 90th | 16.82 | 0.80 | 0.50 | 1.27 |
|  | 95th | 17.79 | 1.82 | 1.15 | 2.89 |
|  | 99th | 18.85 | 8.79 | 5.14 | 15.03 |

